# Trends and determinants of prelacteal feeding practice in rural Bangladesh from 2004 to 2019: A multivariate decomposition analysis

**DOI:** 10.1101/2025.07.11.25331340

**Authors:** Ya Gao, Amanda C. Palmer, Andrew L. Thorne-Lyman, Saijuddin Shaikh, Hasmot Ali, Hannah Tong, Monica M. Pasqualino, Lee S. Wu, Kelsey Alland, Kerry J. Schulze, Alain B. Labrique, Rolf D. Klemm, Parul Christian, Keith P. West

## Abstract

Prelacteal feeding (PLF)—giving infants food or liquid other than breastmilk within the first 3 days of life—remains common and hinders optimal breastfeeding in Bangladesh. This study assessed changes in PLF practices in rural Bangladesh from 2004 to 2019 and examined associate household, maternal, and infant factors. We analyzed data from two cluster-randomized trials in rural northwest Bangladesh (n=16,551; n=4,401). Trained staff collected sociodemographic and birth data through household visits. We used multivariable logistic regression to examine associations between household, maternal, and infant characteristics and PLF and a non-linear approximation of the Oaxaca-Blinder regression decomposition to understand the factors associated with the changing prevalence of PLF. The prevalence of PLF declined from 89% in 2004 to 24% in 2019. Factors associated with PLF shifted over time, particularly household wealth, infant sex, and birth weight. Institutional delivery (OR=0.27; 95% CI 0.22, 0.32 in 2004; OR=0.78; 95% CI 0.61, 1.00 in 2019) and multigravida status (OR=0.68; 95% CI 0.58, 0.79 in 2004; OR=0.73; 95% CI 0.58, 0.93 in 2019) were consistently associated with reduced odds of PLF across cohorts in the multivariable analysis. The decomposition analysis based on the two trials indicated that changes in prevalence of the covariates explained 15% of the decrease in prevalence of PLF, primarily accounted for by increases in health facility deliveries (86%), increases in infant birth weight (13%), and increasing gravidity (12%). 85% of the change remains unexplained by the measured variables. The prevalence of PLF declined considerably in rural Bangladesh over the 15-year period. There are shifts in factors associated with PLF overtime. Improvements in socio-demographic factors played a modest but meaningful role in reducing PLF. However the majority of the reduction remains unexplained by the measured variables. Further research is needed to identify other potential drivers for changes in the prevalence of PLF.

## Introduction

Breastfeeding provides benefits to the immediate health and survival of infants and young children, particularly through its protection against infectious diseases (1, 2). Breastfeeding may also have longer-term benefits, including improved cognitive development (3), as well as reduced risks of overweight and obesity (4–7), hypertension, elevated serum cholesterol, and type 2 diabetes (3, 4) later in life. Given the benefits of breastfeeding, the World Health Organization and United Nations Children’s Fund recommend early initiation of breastfeeding within one hour of birth and exclusive breastfeeding, defined as feeding only breast milk without other liquids or solids, for infants less than 6 months (8). There have been improvements in breastfeeding practices over time, but the latest prevalence of exclusive breastfeeding in Bangladesh (62.6% in 2019) is still short of the World Health Assembly goal of at least 70% by 2030 (9).

One of the barriers for optimal breastfeeding practices is prelacteal feeds, i.e., any fluid or solid other than breastmilk fed before the establishment of breastfeeding, usually within the first 3 days of life. Prelacteal feeding (PLF) is a common practice in South Asia due to ethnic and cultural beliefs (10). The use of honey or sugar-sweetened water has been reported in Bangladesh, as people believe that these sweet prelacteal feeds can clear the voice, prevent infant from catching a common cold, and bless the child with a charming personality in the future (11). Besides cultural beliefs, the perception of insufficient milk production, sickness or unconsciousness after delivery, and stopping infants from crying were also reported reasons for administering PLF from Bangladeshi mothers (11). PLF may delay the initiation of breastfeeding, potentially reducing exposure to colostrum (12, 13), or cause the early cessation of breastfeeding (14–16). Prelacteal foods may also expose vulnerable newborns to pathogens or chemicals, but the longer-term impacts on health and nutritional status of these practices are not well studied(17).

According to the Bangladesh Demographic and Health Survey (DHS), there was a decreasing trend of PLF prevalence at the national level from 62% in 2007 (18) to 24% in 2019 (19). However, the factors that contributed to this trend have not been explored to date. Furthermore, data collection on PLF in the DHS and other large surveys often requires two to five years of recall to get a sufficient sample size. This may introduce recall bias, particularly because PLF often occurs over a period of just a few days (20, 21). This problem can be avoided by collecting data on PLF soon after delivery. Identifying factors associated with changes in the prevalence of PLF overtime may also lead to the development of sustainable interventions and effective recommendations on breastfeeding policies.

To the best of our knowledge, no studies have explored the determinants of the trend in prevalence of PLF over time in Bangladesh. We had the opportunity to analyze data collected in northwest Bangladesh at a site that was considered largely regionally representative and has hosted a variety of trials to assess pregnancy outcomes, such that the questions about PLF could be posed to women shortly after birth. The primary objective of this study is to examine trends in prelacteal feeding (PLF) practices in northwest Bangladesh between 2004 and 2019. Specifically, we aim to (1) quantify changes in the prevalence of PLF over time, (2) identify the socio-demographic factors associated with these changes, and (3) use decomposition analysis to determine the contributions of various explanatory factors, such as maternal education, place of delivery, and birth order, to the observed trends. By addressing these objectives, this study seeks to inform future interventions aimed at reducing harmful feeding practices and promoting optimal breastfeeding

## Materials and Methods

### Study Site Description

The JiVitA project operates in a rural area, spanning 435 km^2^ across 19 unions of the northwest districts of Gaibandha and Rangpur with a population of 650,000 (22). The site has hosted several cluster-randomized controlled trials and observational studies related to maternal and child health (23–25).

### Study Design and Population

The first trial, hereafter referred to as JiVitA-1, was a double-masked, cluster-randomized, placebo-controlled trial assessing the efficacy of maternal vitamin A or beta carotene supplementation in reducing pregnancy-related and infant mortality (23). A cohort of the JiVitA-1 trial (January 2004 to December 2006) that contributed to assessing the effect of supplementing newborns with 50,000 IU of vitamin in reducing all-cause infant mortality through 24 weeks of age was included in the analysis (24). The second trial, hereafter referred to as mCARE-II, was a cluster-randomized controlled trial testing a digital health intervention to improve coverage of antenatal and postnatal care. A cohort of the mCARE-II trial (September 2018 to July 2019) that contributed to future infant growth assessment was included in the analysis (26). All studies identified and recruited participants through a five-weekly pregnancy surveillance system that has been in place across the study areas since its establishment. All women within the study areas who self-reported their pregnancy based on a documented missed menstrual cycle and a positive pregnancy urine test were consented for enrollment and then received follow-up visits from JiVitA research workers throughout their pregnancy and the postpartum period. The details of these trials have been published elsewhere (22–24).

### Data Collection

In both studies, trained field workers collected data on household socioeconomic status and maternal demographic characteristics at the time of enrollment. Upon receiving notification of an infant’s birth, field workers visited the home within 72 hours postpartum, collecting detailed data on breastfeeding initiation and any foods or liquids other than own mother’s breast milk provided to the infant, as well as characteristics surrounding the birth environment, gestational age at birth, infant sex, and other demographic characteristics.

Information on PLF practices in both studies was collected through structured interviews by field interviewers. In the first trial, field interviewers administered the interview as soon as possible after birth. Mothers were asked “What food, other than the mother’s breast milk has the baby been fed? Note: Only include foods given within the first 3 days after birth.” The recorded responses included nothing offered or one or more of the common feeds (cow/goat/sheep/buffalo milk; water; drops; honeys; other mother’s milk; sugar water/misri water; oil, or other). In the second trial, field interviewers visited the households of consented mothers within 72 hours postpartum to conduct the interview. Mothers were asked about time intervals following birth. Specifically, mothers were asked: “Was the baby fed other mother’s breast milk or anything than own mother’s breastmilk in the [first 30 minutes, second 30 minutes, 2nd hour, 3rd hour, 4th hour, 5th hour, 6th hour, 7th-12th hours, 13th-24th hours, remaining hours until upcoming 6 am after completion of 24-hour, entire day 3 and night]?” For each time interval, if the mother responded yes, data on feeding from other mother’s milk or specific foods from a list of common non-breast milk feeds (honey, water, animal milk, formula, sugar/sugar candy water, any types of drops, powdered/condensed milk, and others) were also collected. “Drops” in Bangladesh consisted of a wide range of items. Previous studies found drops to include homeopathic supplements and broad-spectrum antibiotics, concentrated vitamin supplements, and purportedly sterile solutions of sucrose, glucose, dextrose, or saline (27).

### Statistical Analysis

For this analysis, we included consented women with singleton live births. To reduce the likelihood of recall bias, we also restricted our analysis to interviews conducted within 30 days postpartum. PLF was dichotomized as “yes” or “no”. For the first trial, yes was defined as any responses other than “nothing offered”. For the second trial, yes was defined as one or more yes responses to the question within the first 3 days after birth.

The potential determinants of PLF used for analyses were identified after consulting similar literature (27, 28). The selected variables were infant gestational age at birth, infant sex, type of delivery, birth location, infant birth weight, maternal age, maternal literacy, maternal education, maternal gravidity, participation in any micro-credit program, and household socioeconomic status. A Living Standards Index (LSI) was constructed using principal component analysis of data on 20 socioeconomic factors, including household assets and house construction materials (29). LSI was then categorized into five wealth quintiles, with the highest quintile corresponding to the wealthiest group and the lowest quintile representing the poorest. Five categories of birth locations were combined into two categories. One category included home and enroute/other. The other category, institutional deliveries, included family welfare visitor’s house, health or welfare center, and hospital/clinic/medical college.

Bivariate and multivariable logistic regression models were used to examine the associations between different demographic characteristics and PLF status. A value of P < 0.05 was considered statistically significant. None of the covariates included in the multivariate models had variance inflation factors higher than 3 or tolerance values less than 0.1, suggesting that there was no multicollinearity among covariates in the final models.

To examine the extent to which the differences in the prevalence of PLF over the 15-year period from 2004 to 2019 were due to changes in maternal and infant demographic characteristics, a non-linear approximation of the Oaxaca-Blinder regression decomposition technique was used (30). We used Stata’s *mvdcmp* command and included a full set of maternal and infant demographic variables related to prelacteal feeding in the decomposition model. Data management and statistical analyses were conducted in Stata Version 15.1.21.

### Ethics Statement

The protocols for the JiVitA-1 and mCARE-II trial (IRB No. 00006469, approved on August 27, 2015) were reviewed and approved by institutional review board at the Bloomberg School of Public Health at Johns Hopkins University and Bangladesh Medical Research Council, Dhaka, Bangladesh. Individual participants provided written informed consent.

## Results

A total of 16,551 infants enrolled in the JiVitA-1 cluster-randomized trial from 2004-2006, and 4,401 infants enrolled in the mCARE-II cluster-randomized trial from 2018-2019, were included in the final analysis (**Supplement Figure 1**). Maternal and infant characteristics of the two study cohorts are summarized in **Table 1**. The percentage of women giving birth before the age of 19 years decreased from 41.5% to 26.8% over this period while the literacy rate among women increased from 48.6% to 81.1%. The percentage of premature births dropped from 26.8% to 17.7%. Health Facility deliveries increased from 6.1% to 40.1%, including an increase in delivery at hospital, clinic, or medical college (2.5% to 28.1%). Cesarean delivery rose markedly from 1.9% to 25.1% in the study area.

**Table 1:**
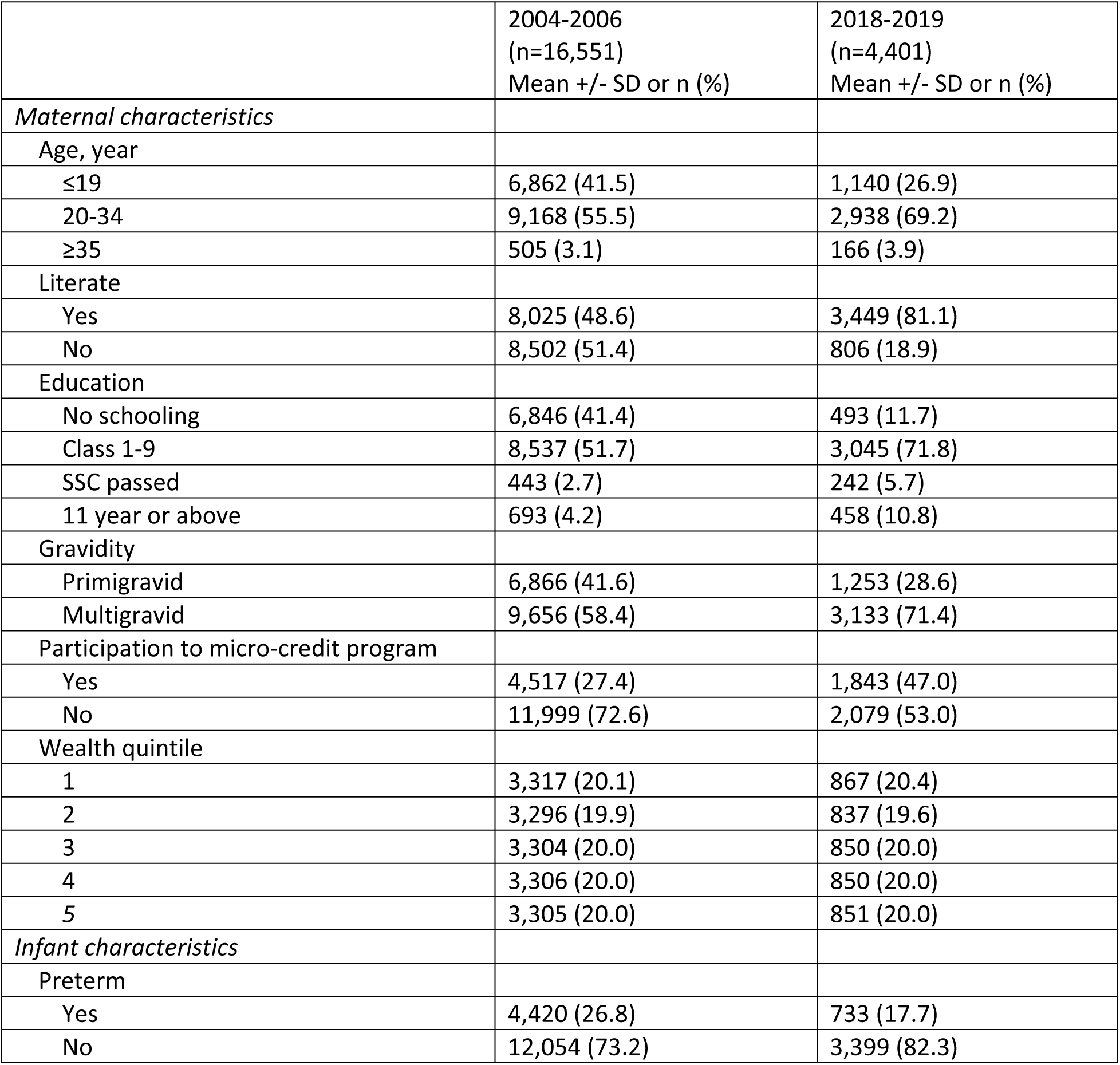
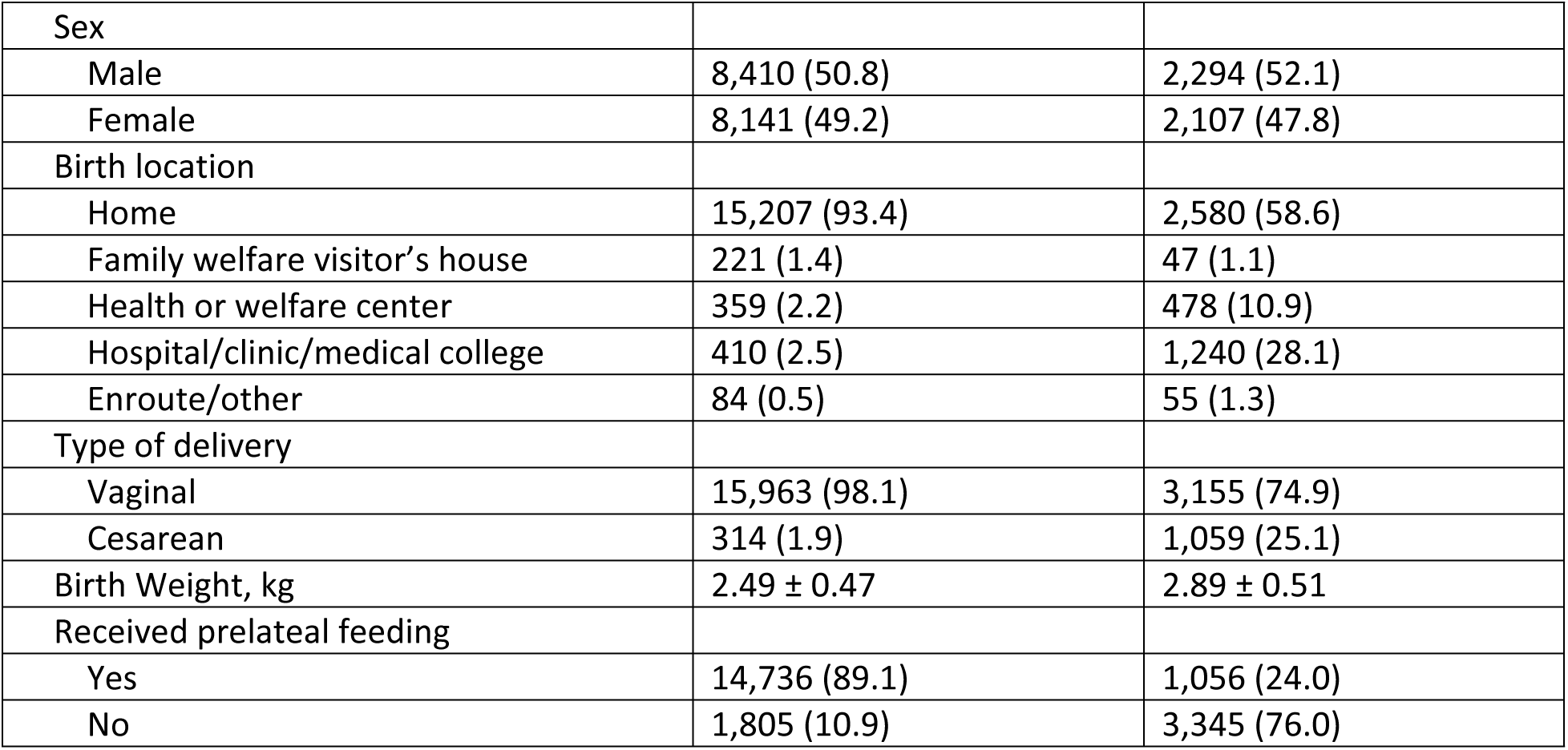
Maternal and infant characteristics of the two study cohorts in rural Bangladesh.

|  | 2004-2006<br>(n=16,551)<br>Mean +/- SD or n (%) | 2018-2019<br>(n=4,401)<br>Mean +/- SD or n (%) |
| --- | --- | --- |
| *Maternal characteristics* |  |  |
| Age, year |  |  |
| ≤19 | 6,862 (41.5) | 1,140 (26.9) |
| 20-34 | 9,168 (55.5) | 2,938 (69.2) |
| ≥35 | 505 (3.1) | 166 (3.9) |
| Literate |  |  |
| Yes | 8,025 (48.6) | 3,449 (81.1) |
| No | 8,502 (51.4) | 806 (18.9) |
| Education |  |  |
| No schooling | 6,846 (41.4) | 493 (11.7) |
| Class 1-9 | 8,537 (51.7) | 3,045 (71.8) |
| SSC passed | 443 (2.7) | 242 (5.7) |
| 11 year or above | 693 (4.2) | 458 (10.8) |
| Gravidity |  |  |
| Primigravid | 6,866 (41.6) | 1,253 (28.6) |
| Multigravid | 9,656 (58.4) | 3,133 (71.4) |
| Participation to micro-credit program |  |  |
| Yes | 4,517 (27.4) | 1,843 (47.0) |
| No | 11,999 (72.6) | 2,079 (53.0) |
| Wealth quintile |  |  |
| 1 | 3,317 (20.1) | 867 (20.4) |
| 2 | 3,296 (19.9) | 837 (19.6) |
| 3 | 3,304 (20.0) | 850 (20.0) |
| 4 | 3,306 (20.0) | 850 (20.0) |
| 5 | 3,305 (20.0) | 851 (20.0) |
| *Infant characteristics* |  |  |
| Preterm |  |  |
| Yes | 4,420 (26.8) | 733 (17.7) |
| No | 12,054 (73.2) | 3,399 (82.3) |
| Sex |  |  |
| Male | 8,410 (50.8) | 2,294 (52.1) |
| Female | 8,141 (49.2) | 2,107 (47.8) |
| Birth location |  |  |
| Home | 15,207 (93.4) | 2,580 (58.6) |
| Family welfare visitor's house | 221 (1.4) | 47 (1.1) |
| Health or welfare center | 359 (2.2) | 478 (10.9) |
| Hospital/clinic/medical college | 410 (2.5) | 1,240 (28.1) |
| Enroute/other | 84 (0.5) | 55 (1.3) |
| Type of delivery |  |  |
| Vaginal | 15,963 (98.1) | 3,155 (74.9) |
| Cesarean | 314 (1.9) | 1,059 (25.1) |
| Birth Weight, kg | 2.49 ± 0.47 | 2.89 ± 0.51 |
| Received prelacteal feeding |  |  |
| Yes | 14,736 (89.1) | 1,056 (24.0) |
| No | 1,805 (10.9) | 3,345 (76.0) |

The prevalence of any form of PLF was 89.1% during the period 2004-2006 (**Table 2**). Among the infants who received PLF, sugar-sweetened water was the most common prelacteal feed (47.2%), followed by animal milk (45.6%), honey (40.9%), and drops (13.4%) (**Figure 1**). In 2018-2019, the prevalence of PLF had dropped to 24.0% and the most common prelacteal feed was animal milk (20.6%), followed by sugar-sweetened water (20.5%), honey (20.5%), and any type of drops (14.4%).

**Figure 1.**
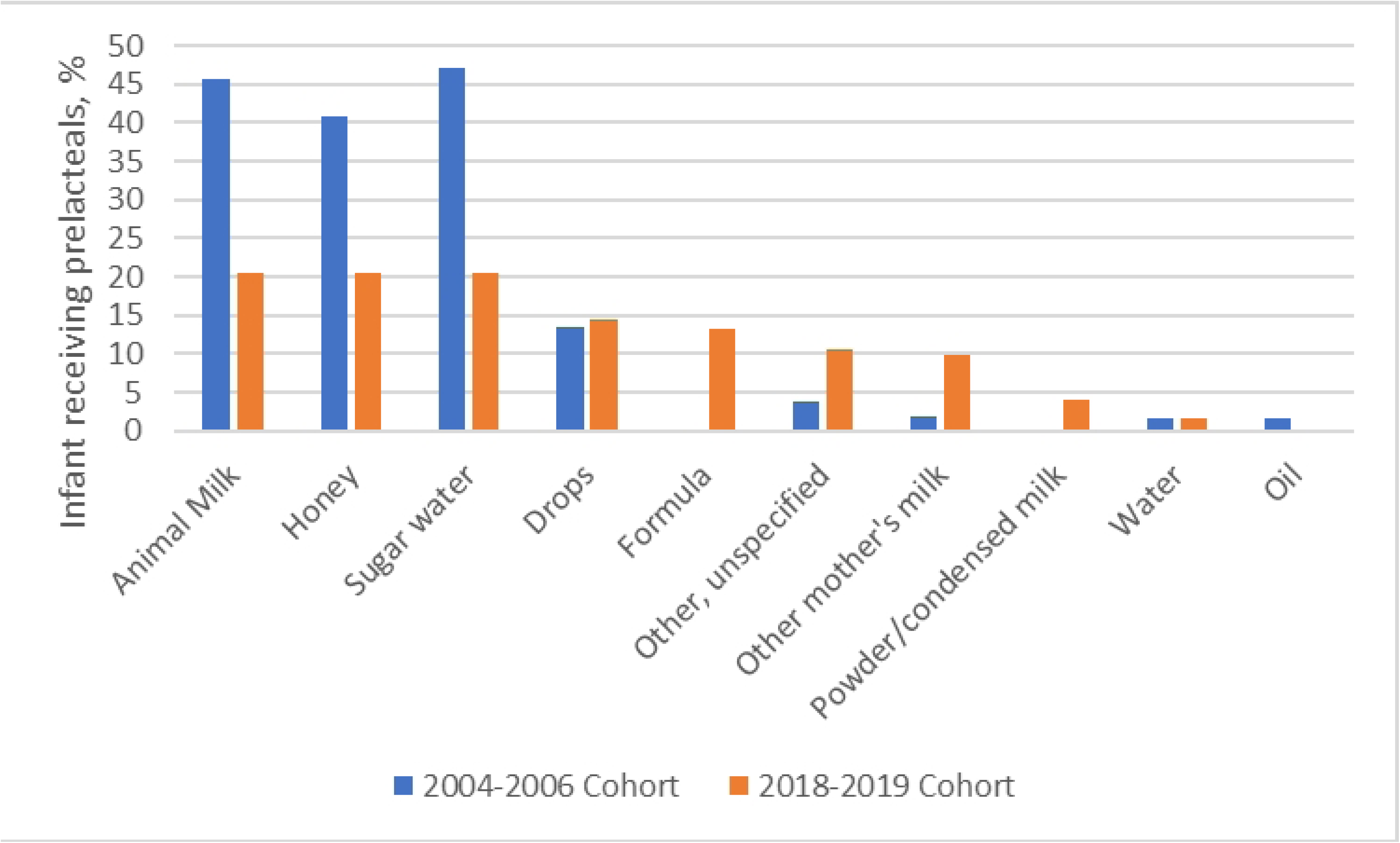
Change in frequency of prelacteal feeding by types in rural Bangladesh from 2004 to 2019. The frequencies are nonexclusive, meaning that it was possible for one woman to feed multiple types of foods. Data about formula was not collected in 2004-2006 cohort and data about oil was not collected in 2018-2019 cohort.

**Table 2.** Maternal and infant characteristics of the two study cohorts in rural Bangladesh by prelacteal feeding status.

|  | 2004 to 2006 cohort |  | 2018 to 2019 cohort |  |
| --- | --- | --- | --- | --- |
|  | Feeding<br>prelacteals<br>(n=14,736) | Not feeding<br>prelacteals<br>(n=1,805) | Feeding<br>prelacteals<br>(n=3,345) | Not feeding<br>prelacteals<br>(n=1,056) |
|  | Mean ± SD<br>or n (%) | Mean ± SD<br>or n (%) | Mean ± SD<br>or n (%) | Mean ± SD<br>or n (%) |
| *Maternal characteristics* |  |  |  |  |
| Age, year |  |  |  |  |
| ≤19 | 6,267 (42.6) | 590 (32.7) | 281 (27.7) | 859 (26.6) |
| 20-34 | 8,003 (54.4) | 1,160 (64.3) | 687 (67.8) | 2,251 (69.7) |
| ≥35 | 450 (3.1) | 55 (3.1) | 46 (4.5) | 120 (3.7) |
| Literate |  |  |  |  |
| Yes | 7,135 (48.5) | 887 (49.2) | 825 (81.3) | 2,624 (81.0) |
| No | 7,580 (51.5) | 915 (50.8) | 190 (18.7) | 616 (19.0) |
| Education |  |  |  |  |
| No schooling | 6,085 (41.4) | 754 (41.9) | 124 (12.3) | 374 (11.6) |
| Class 1-9 | 7,675 (52.2) | 860 (47.7) | 722 (71.4) | 2,323 (71.9) |
| SSC passed | 378 (2.5) | 65 (3.5) | 59 (5.8) | 83 (5.6) |
| 11 year or above | 569 (3.9) | 123 (6.8) | 106 (10.5) | 352 (10.9) |
| Gravidity |  |  |  |  |
| Primigravid | 6,252 (42.5) | 608 (33.8) | 334 (31.7) | 919 (27.6) |
| Multigravid | 8,460 (57.5) | 1,192 (66.2) | 720 (68.3) | 2,413 (72.4) |
| Participation to micro-credit program |  |  |  |  |
| Yes | 3,992 (27.2) | 525 (29.2) | 428 (46.1) | 1,415 (47.3) |
| No | 10,714 (72.8) | 1,275 (70.8) | 500 (53.9) | 1,579 (52.7) |
| Wealth quintile |  |  |  |  |
| 1 | 2,940 (20.0) | 375 (20.8) | 193 (19.0) | 674 (20.8) |
| 2 | 2,964 (20.1) | 329 (18.2) | 199 (19.6) | 638 (19.7) |
| 3 | 2,973 (20.2) | 331 (18.4) | 190 (18.7) | 660 (20.4) |
| 4 | 2,958 (20.1) | 347 (19.2) | 213 (21.0) | 637 (19.6) |
| 5 | 2,881 (19.6) | 421 (23.4) | 220 (21.7) | 631 (19.5) |
| *Infant characteristics* |  |  |  |  |
| Preterm |  |  |  |  |
| Yes | 3,944 (26.9) | 476 (26.5) | 174 (17.5) | 559 (17.8) |
| No | 10,725 (73.1) | 1,319 (73.5) | 819 (82.5) | 2,580 (82.2) |
| Sex |  |  |  |  |
| Male | 7,508 (51.0) | 898 (49.8) | 585 (55.4) | 1,710 (51.1) |
| Female | 7,228 (49.0) | 907 (50.2) | 471 (44.6) | 1,635 (48.9) |
| Birth location |  |  |  |  |
| Home† | 13,793 (95.2) | 1,491 (84.0) | 660 (62.6) | 1,975 (59.0) |
| Health Facility‡ | 702 (4.8) | 285 (11.5) | 395 (55.0) | 1,370 (41.0) |
| Type of delivery |  |  |  |  |
| Vaginal | 14,279 (98.5) | 1,675 (94.3) | 773 (76.3) | 2,382 (74.4) |
| Cesarean | 211 (1.5) | 102 (5.7) | 240 (23.7) | 819 (25.6) |
| Birth Weight, kg | 2.48 ± 0.46 | 2.56 ± 0.48 | 2.88 ± 0.52 | 2.89 ± 0.51 |

†Includes home and enroute/other, number of births happened enroute/other is much smaller than number of births happened at home, See Table 1.
‡Includes family welfare visitor's houses; or health or welfare center; or Hospital/clinic/medical college.

The bivariate and multivariate analyses of the determinants of PLF among mothers and infants enrolled in the 2004-2006 cohort are shown in **Table 3**. In bivariate analyses, PLF was associated with younger maternal age, lower maternal education, maternal primigravity, vaginal delivery, home delivery, and lower infant birth weight. In the multivariate regression model, maternal multigravidity (OR=0.68; 95% CI 0.58, 0.79); health facility deliveries (OR=0.27; 95% CI 0.22, 0.32); and higher infant birth weight (OR=0.81; 95% CI 0.72, 0.91) reduced the odds of PLF.

**Table 3.** Odds of prelacteal feeding based on characteristics of women and infants of the two study cohorts.

|  | 2004 to 2006 cohort |  | 2018 to 2019 cohort |  |
| --- | --- | --- | --- | --- |
|  | Feeding<br>prelacteals<br>(crude) | Feeding<br>prelacteals<br>(adjusted) | Feeding<br>prelacteals<br>(crude) | Feeding<br>prelacteals<br>(adjusted) |
|  | OR (95% CI) | OR (95% CI) | OR (95% CI) | OR (95% CI) |
| *Maternal characteristics* |  |  |  |  |
| Age, year |  |  |  |  |
| ≤19 | 1.0 (ref.) | 1.0 (ref.) | 1.0 (ref.) | 1.0 (ref.) |
| 20-34 | 0.65 (0.59-0.72)* | 0.89 (0.76-1.03) | 0.93 (0.80-1.09) | 1.08 (0.86, 1.36) |
| ≥35 | 0.77 (0.57-1.03) | 1.04 (0.75-1.44) | 1.17 (0.81-1.69) | 1.40 (0.91, 2.15) |
| Literate |  |  |  |  |
| Yes | 1.0 (ref.) | 1.0 (ref.) | 1.0 (ref.) | 1.0 (ref.) |
| No | 1.03 (0.93-1.14) | 1.06 (0.88, 1.27) | 0.98 (0.82-1.43) | 0.92 (0.68, 1.24) |
| Education |  |  |  |  |
| No schooling | 1.0 (ref.) | 1.0 (ref.) | 1.0 (ref.) | 1.0 (ref.) |
| Class 1-9 | 1.11 (1.00-1.23) | 1.06 (0.89-1.27) | 0.94 (0.75-1.17) | 0.83 (0.59, 1.18) |
| SSC passed | 0.72 (0.55-0.95)* | 0.98 (0.69-1.38) | 0.97 (0.68-1.39) | 0.79 (0.47, 1.30) |
| 11 year or above | 0.57 (0.47-0.71)* | 0.94 (0.70-1.27) | 0.91 (0.67-1.22) | 0.72 (0.45, 1.13) |
| Gravidity |  |  |  |  |
| Primigravid | 1.0 (ref.) | 1.0 (ref.) | 1.0 (ref.) | 1.0 (ref.) |
| Multigravid | 0.69 (0.62-0.77)* | 0.68 (0.58-0.79)* | 0.82 (0.71-0.95)* | 0.73 (0.58-0.93)* |
| Participation to micro-credit program |  |  |  |  |
| Yes | 1.0 (ref.) | 1.0 (ref.) | 1.0 (ref.) | 1.0 (ref.) |
| No | 1.11 (0.99-1.23) | 1.06 (0.95-1.19) | 1.05 (0.90-1.21) | 0.96 (0.81, 1.13) |
| Wealth quintile |  |  |  |  |
| 1 | 1.0 (ref.) | 1.0 (ref.) | 1.0 (ref.) | 1.0 (ref.) |
| 2 | 1.15 (0.98-1.34) | 1.17 (1.00-1.38) | 1.09 (0.87-1.37) | 1.11 (0.87-1.41) |
| 3 | 1.15 (0.98-1.34) | 1.17 (0.99-1.37) | 1.01 (0.80-1.26) | 1.03 (0.81-1.33) |
| 4 | 1.09 (0.93-1.27) | 1.12 (0.95-1.33) | 1.17 (0.93-1.46) | 1.09 (0.84-1.41) |
| 5 | 0.87 (0.75-1.01) | 1.01 (0.84-1.21) | 1.22 (0.98-1.52) | 1.36 (1.03-1.81)* |
| *Infant characteristics* |  |  |  |  |
| Preterm |  |  |  |  |
| Yes | 1.0 (ref.) | 1.0 (ref.) | 1.0 (ref.) | 1.0 (ref.) |
| No | 0.98 (0.88-1.10) | 1.06 (0.94, 1.19) | 1.01 (0.85-1.23) | 1.00 (0.82, 1.23) |
| Sex |  |  |  |  |
| Male | 1.0 (ref.) | 1.0 (ref.) | 1.0 (ref.) | 1.0 (ref.) |
| Female | 0.95 (0.86-1.05) | 0.92 (0.83, 1.02) | 0.84 (0.73-0.97)* | 0.83 (0.71-0.97)* |
| Birth location |  |  |  |  |
| Home† | 1.0 (ref.) | 1.0 (ref.) | 1.0 (ref.) | 1.0 (ref.) |
| Health Facility‡ | 0.27 (0.23, 0.31)* | 0.27 (0.22, 0.32)* | 0.86 (0.75, 0.99)* | 0.78 (0.61, 1.00)* |
| Type of delivery |  |  |  |  |
| Vaginal | 1.0 (ref.) | 1.0 (ref.) | 1.0 (ref.) | 1.0 (ref.) |
| Cesarean | 0.24 (0.19-0.31)* | 0.89 (0.65-1.19) | 0.90 (0.77-1.07) | 1.05 (0.79, 1.39) |
| Birth Weight, kg | 0.69 (0.62-0.76)* | 0.81 (0.72-0.91)* | 0.97 (0.84-1.10) | 1.01 (0.86, 1.18) |

\*Factor significantly associated with prelacteal feeding compared to its reference group, P<0.05.
†Includes home and enroute/other, number of births happened enroute/other is much smaller than number of births happened at home, See Table 1.
‡Includes family welfare visitor's houses; or health or welfare center; or Hospital/clinic/medical college.

The bivariate and multivariate analyses of the determinants of PLF among mothers and infants enrolled in the 2018-2019 cohort are shown in **Table 3**. Maternal primigravity, male infant sex, and home delivery were associated with higher odds of PLF in bivariate analyses. In the multivariate regression model, maternal multigravidity (OR=0.73; 95% CI 0.58, 0.93); female infant sex (OR=0.83; 95% CI 0.71, 0.97); and health facility deliveries (OR=0.78; 95% CI 0.61, 1.00) reduced the odds of PLF. When compared to the lowest wealth quintile, the children from wealthiest quintile household had higher odds of PLF (OR=1.36; 95% CI 1.03, 1.51).

Maternal age, literacy, gravidity, participation in micro-credit program, infant sex, birth weight, birth location, and type of delivery were included in the Oaxaca-Blinder decomposition model to examine the extent to which changes in these factors contributed to the change in PLF prevalence overtime in rural Bangladesh. About 15% of the decrease in PLF prevalence from 2004 to 2019 was explained by the changes in the maternal and infant demographic characteristics (**Table 4**). Among the explained components, the increase in health facility deliveries contributed most to the reduction in PLF (62%), followed by increase in multigravida (12%), increased average infant birth weight (13%), increase in cesarean delivery (5%), increased maternal age at birth (5%), and improved maternal literacy (2%) (**Figure 2**). The disaggregated results for the Oaxaca-Blinder decomposition of change in PLF between 2004 and 2019 were reported in **Supplementary Table 1**.

**Figure 2.**
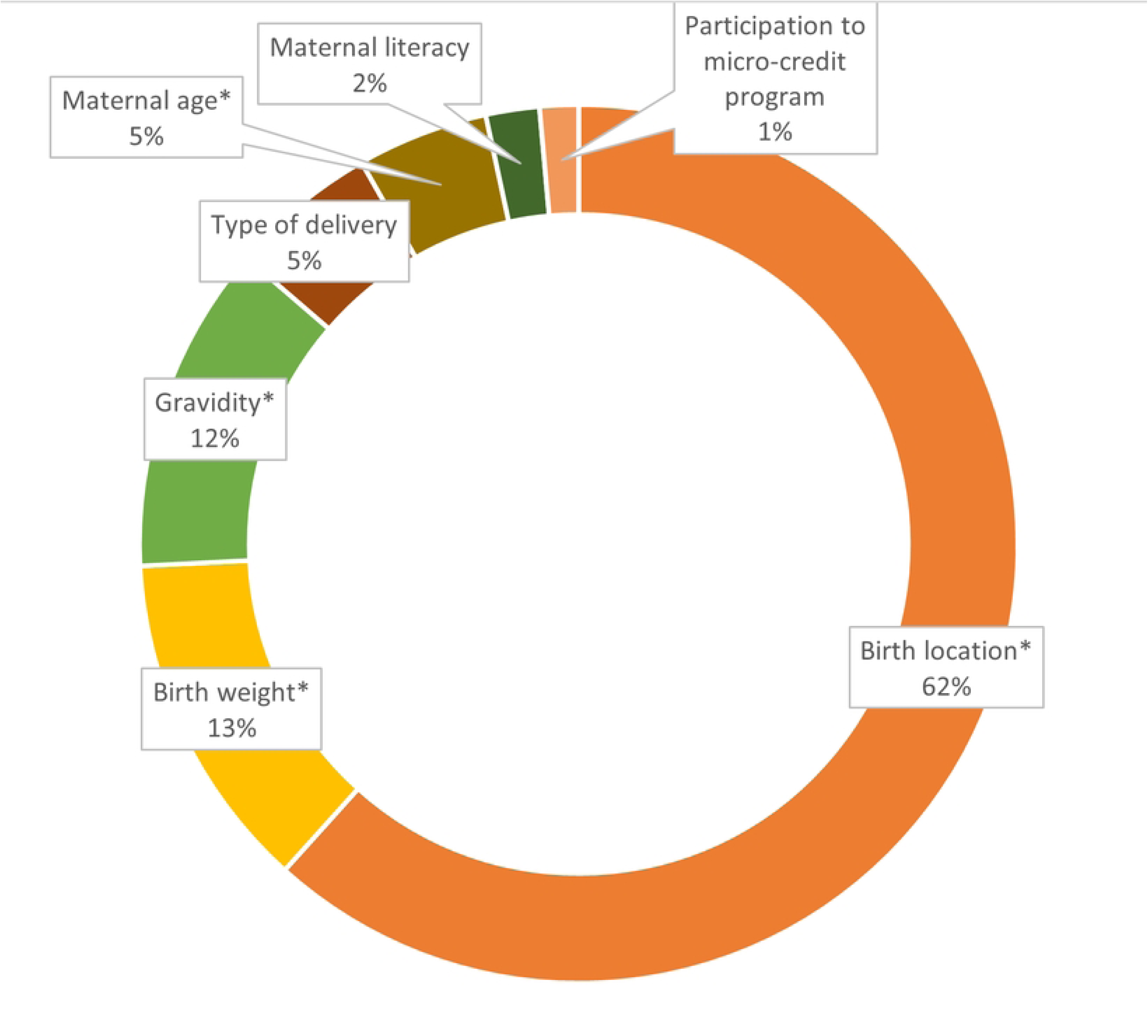
Contributions to PLF prevalence reduction by maternal and infant characteristics. *P value <0.05.

**Table 4.**
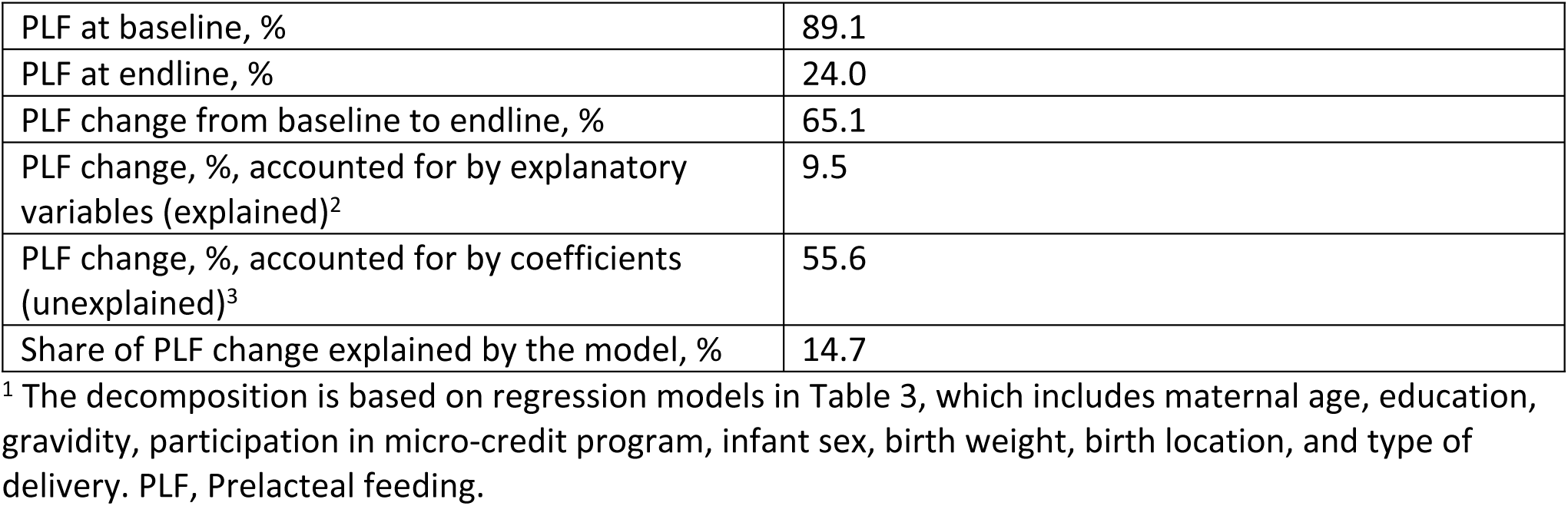
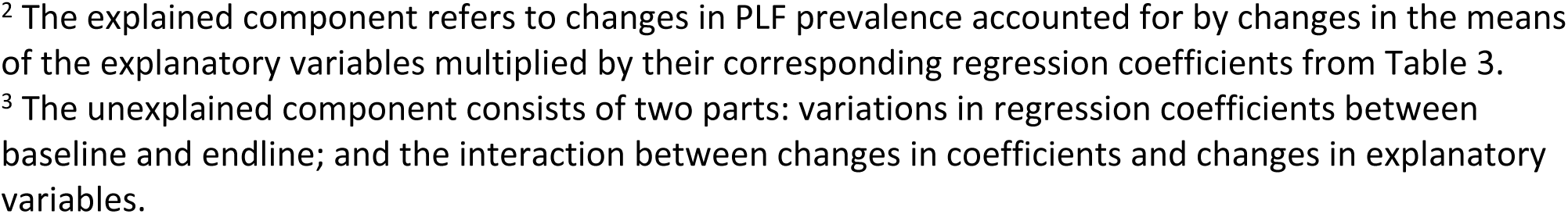
Summary Results for the Oaxaca-Blinder decomposition of change in PLF prevalence between 2004 and 2019 in Bangladesh^1^.

## Discussion

The prevalence of PLF declined over the 15-year observation period from 89% in 2004 to 24% in 2019 in our study area. Institutional delivery and multigravida status were consistently associated with reduced odds of PLF across cohorts; however, the importance of institutional delivery decreased in the more recent cohort (2018-2019). Infant weight was no longer a significant determinant, while the highest wealth category became a significant predictor of PLF in the more recent cohort. The sex of the infant played an increasingly important role in the more recent cohort, with female infants being less likely to receive PLF in 2018-2019. The finding suggests shifts in factors associated with PLF over time, particularly related to household wealth, infant sex, and birth weight. Over the 15-year period, the changes in prevalence of the covariates explained 15% of the decrease in prevalence of PLF, primarily accounted for by increases in health facility deliveries, increasing gravidity, an increase in infant birth weight, and increasing maternal age.

Sugar water, animal milk, honey, and drops remain the most commonly fed prelacteals in this rural Bangladeshi setting over the 15-year period. Our results are in alignment with a cross-sectional study carried out in the Tangail district in rural Bangladesh, where they found infants were most commonly fed sugar water, followed by drops, infant formula, honey, and other milk than breastmilk (31). The preference of sugar water and honey conveys the cultural beliefs and ritual practices around giving something sweet to newborns (32).

Despite the considerable decline in PLF prevalence over the past 15 years, 1 in 4 women still fed their newborns prelacteals in this rural Bangladeshi setting. The provision of prelacteal feeds, by definition, disrupts exclusive breastfeeding and may have displaced colostrum. As the “first milk” produced between birth through the first 5 days of lactation, colostrum has been found to have a variety of nutritional and immunological benefits to neonates (33). Moreover, in rural settings with poor hygiene, the preparation of prelacteal feeds may potentially introduce harmful substances, such as heavy metals and pathogens in contaminated water. The provision of these feeds to neonates may increase the risk of infectious diseases such as diarrhea and pneumonia, as well as other acute infections and allergies, compromising infant growth and development. However, there are no conclusive findings on the impact of PLF on infant growth (17, 34, 35).

In both study cohorts, women giving birth at home were at a higher risk of feeding their newborns prelacteals than women who gave birth at health centers of medical institutions. Similar associations between home delivery and PLF have also been reported in India and Pakistan (36, 37). In addition, the results of decomposition analysis indicated the increase in health facility delivery, from 6.6% to 41.4%, to be the primary driver of the change in PLF, explaining 9.1% of the reduction in PLF prevalence over the past 15 years. Another decomposition analysis study in Ethiopia found that increase in health facility delivery, from 6.4% to 35.6%, explained 7.8% of the reduction in PLF (38). Mothers giving birth at home were more likely to be influenced by their family, friends, and unskilled birth attendants who based their advice on cultural beliefs or personal experience, which may facilitate the practice of PLF. Attending an institutional delivery would have exposed mothers to infant and young child feeding education and immediate postnatal care, such as the encouragement of early initiation of breastfeeding, which may reduce their tendency to feed prelacteals (39, 40). The government of Bangladesh has taken steps to encourage institutional delivery (41). A pilot maternity voucher scheme, providing monetary incentive for attending antenatal care and delivery at public or private facility, or at home with a skilled birth attendant, reached more than 10 million people (42).

Multiparous mothers have been found to be less likely to practice PLF in this rural Bangladeshi setting over the 15-year period. Similar findings have been reported by two studies in Nepal, that first time mothers tended to have a higher likelihood of giving prelacteals (43, 44). It is likely that first time mothers who had no previous child feeding experiences were more likely to be guided by advice from family members who encouraged PLF (45). In addition, prior studies have also revealed that multiparous mothers were more likely to initiate breastfeeding early after delivery, and those who had prior breastfeeding experiences would maintain breastfeeding for a longer duration compared with first time mothers (46). In the decomposition model, the increased gravidity over time also explained 1.8% of the reduction in PLF prevalence. It is important to note that the 2004 cohort recruited more newly married women to the surveillance activities to ensure that enough new pregnancies were captured, thereby the 2004 cohort is not representative of the pregnant women at the JiVitA site. The 2019 cohort provides a better representation of pregnant women at the JiVitA site. The contribution to the change might be smaller had the older cohort provided a better snapshot of the pregnant women in the study site.

Only 9.5% of the 65.1% reduction in PLF is explained by observable changes in factors such as birth location, gravidity, birth weight, and maternal age. This suggests that improvements in these socio-demographic factors played a modest by meaningful role in reducing PLF. But the majority (55.6%) of the reduction in PLF remains unexplained by the measured variables included in the analysis. The scaling up of infant and young child feeding education at health care facilities, as well as increased access to various breastfeeding promotion campaigns may have contributed to a greater extent, the decline in PLF, as evidenced by decline in prevalence of PLF at national level from 62% in 2007 to 29% in 2018 (18, 47). A large-scale program to improve infant and young child feeding practices was implemented in Bangladesh, from 2010 to 2014, covering 50 rural sub-districts, through the existing national Essential Health Care program (48). The at-scale program lowered the use of PLF through interpersonal counseling, mass media, community mobilization, and policy advocacy (48). Since the program covered JiVitA site, women residing in JiVitA site might be indirectly influenced. However limited data on breastfeeding advocation have been collected in this setting to be able to quantify its effects. Our current analysis adds to the literature on the changes, as well as drivers of changes, in PLF in rural Bangladesh over the past 15 years. We had the advantage of having data on PLF collected at similar time points between the two studies that were conducted 15 years apart, which enabled the comparison of data over time. Meanwhile, information on PLF was captured soon after birth, which substantially reduced the likelihood of recall bias.

While this study provides important insights into trends in PLF practices, there are several limitations that must be acknowledged. First, the design of the two studies were slightly different. JiVitA-1 had the aim of determining the efficacy of providing an oral supplement containing the weekly equivalent of an RDA of vitamin A from the first trimester of pregnancy through 12weeks (84thday) after pregnancy termination, in reducing all-cause maternal mortality. Newly married women were added to the surveillance activities to ensure that sufficient sample size was included, thereby boosting the number of primiparous women. mCARE-II only identified and recruited participants through the pregnancy surveillance system, providing a snapshot of all pregnant women at the site. The differences in design elicited a different balance of parity, resulting in increased parity in more recent years despite the overall reduction in total fertility rate within Bangladesh (18, 47). Second, the cross-sectional nature of the data limits our ability to establish causality between the observed socio-demographic changes and PLF. Additionally, shifts in cultural practices, healthcare policies, and public health messaging during the 15-year period could have influenced both the prevalence of PLF and associated factors, but these contextual changes were not explicitly captured in the data.

Furthermore, there were differences in survey questions between the two periods. We cannot be certain if the same responses would be elicited if ask two different ways. The differences might be a potential source of bias. Some types of prelacteal feeds were not consistently recorded across both survey periods, limiting our ability to conduct a direct comparison of feeding practices over time. However, we had collected data on six different types of prelacteal foods in both studies, which allowed us to characterize changes in common prelacteal practices over time. But we were not powered to look at the trend in types of PLF, for example milk-based vs water-based, over time. Milk-based PLF were more common in higher-middle-income countries, whereas water-based PLF were more common in low-income countries (9).

## Conclusions

The prevalence of PLF declined considerably by 65% in rural Bangladesh over the 15-year period from 2004 to 2019. Among women practicing PLF, sugar sweetened water, animal milk, honey, and drops remained to be the most frequently fed prelacteals over time. In multivariate models, PLF, in this rural Bangladeshi setting, was significantly associated with lower infant birth weight, home delivery, male infant gender, and maternal primigravidity. Approximately 15% of the reduction in PLF between 2004-2006 and 2018-2019 can be attributed to changes in socio-demographic characteristics, most notably birth weight and institutional delivery rates. 85% of the change remains unexplained by the measured variables. This suggests that other factors, such as shifts in cultural norms, improvements in public health campaigns promoting exclusive breastfeeding, or changes in healthcare delivery, may have contributed to the decline in PLF. Future research should explore these potential influences to gain a more comprehensive understanding of what drives reductions in PLF. Additionally, it is worth considering whether policy changes, such as increased access to skilled birth attendants or the introduction of new breastfeeding promotion programs, may have indirectly influenced mother’s feeding practices.

## Data Availability

Data described in the manuscript, code book, and analytic code will be made available upon request pending application and approval.

## Acknowledgements

We are grateful to the mothers, infants, and their household members for participation in the study. We also thank the many JiVitA field workers and the management team for their support and assistance with recruitment and data collection.

## Supporting information

**S1**. Supplementary figure and table

